# Ethnicity, Household Composition and COVID-19 Mortality: A National Linked Data Study

**DOI:** 10.1101/2020.11.27.20238147

**Authors:** Vahé Nafilyan, Nazrul Islam, Daniel Ayoubkhani, Clare Gilles, Srinivasa Vittal Katikireddi, Rohini Mathur, Annabel Summerfield, Karen Tingay, Miqdad Asaria, Ann John, Peter Goldblatt, Amitava Banerjee, Myer Glickman, Kamlesh Khunti

## Abstract

**Background:** Ethnic minorities have experienced disproportionate COVID-19 mortality rates. We estimated associations between household composition and COVID-19 mortality in older adults (≥65 years) using a newly linked census-based dataset, and investigated whether living in a multi-generational household explained some of the elevated COVID-19 mortality amongst ethnic minority groups.

**Methods:** Using retrospective data from the 2011 Census linked to Hospital Episode Statistics (2017-2019) and death registration data (up to 27^th^ July 2020), we followed adults aged 65 years or over living in private households in England from 2 March 2020 until 27 July 2020 (n=10,078,568). We estimated hazard ratios (HRs) for COVID-19 death for people living in a multi-generational household compared with people living with another older adult, adjusting for geographical factors, socio-economic characteristics and pre-pandemic health. We conducted a causal mediation analysis to estimate the proportion of ethnic inequalities explained by living in a multi-generational household.

**Results:** Living in a multi-generational household was associated with an increased risk of COVID-19 death. After adjusting for confounding factors, the HRs for living in a multi-generational household with dependent children were 1.13 [95% confidence interval 1.01-1.27] and 1.17 [1.01-1.35] for older males and females. The HRs for living in a multi-generational household without dependent children were 1.03 [0.97 - 1.09] for older males and 1.22 [1.12 - 1.32] for older females. Living in a multi-generational household explained between 10% and 15% of the elevated risk of COVID-19 death among older females from South Asian background, but very little for South Asian males or people in other ethnic minority groups.

**Conclusion:** Older adults living with younger people are at increased risk of COVID-19 mortality, and this is a notable contributing factor to the excess risk experienced by older South Asian females compared to White females. Relevant public health interventions should be directed at communities where such multi-generational households are highly prevalent.

**Funding:** This research was funded by the Office for National Statistics.

**Research in context:** *Evidence before this study:* A systematic review by Sze and colleagues demonstrated that people of ethnic minority background in the UK and the USA have been disproportionately affected by the Coronavirus disease 2019 (COVID-19), compared to White populations. We reviewed all papers included within the above systematic review to identify studies which empirically explored potential mediating pathways underpinning ethnic inequalities in COVID-19. In addition, we searched *Pubmed* for studies related to the association between household composition and COVID-19 risk, using the terms ‘household’, ‘COVID-19’ and ‘mortality’, ‘death’ or ‘infection’ on 1 December 2020. Previous research has demonstrated that household size is associated with COVID-19 risk, but there is a lack of studies based on nationally representative individual records that examine the links between household composition and COVID-19 risk. In addition, no study has focused on multigenerational households. Whilst several studies have examined whether socio-demographic and economic factors are driving ethnic inequalities in COVID-19, no study has sought to explicitly quantify the contribution of household composition to the elevated risk of COVID-19 mortality in ethnic minority groups.

*Added value of this study:* Using retrospective data from the 2011 Census linked to Hospital Episode Statistics and death registration data for England, we examined the relationship between household composition and COVID-19 mortality risk amongst older adults (≥65 years). Living in a multi-generational household was associated with an increased risk of COVID-19 death. The adjusted Hazard Ratios (HRs) for living in a multi-generational household with dependent children were 1.13 [95% confidence interval 1.01-1.27] and 1.17 [1.01-1.35] for older males and females. The HRs for living in a multi-generational household without dependent children were 1.03 [0.97 - 1.09] for older males and 1.22 [1.12 - 1.32] for older females. Using a causal mediation approach, we estimated whether the higher propensity to live in multi-generational household amongst ethnic minority groups contributed to their raised risk of COVID-19. We found that living in a multi-generational household explained between 10% and 15% of the elevated risk of COVID-19 death among older females from South Asian background, but very little for South Asian males or people in other ethnic minority groups.

*Implications of all the available evidence:* Living in a large, multi-generational household is associated with an increased risk of COVID-19 infection and death. The increase in risk appears greater for older women than men living in a multi-generational household, and this is particularly the case when living with dependent children. It explains some of the excess COVID-19 mortality risk for females of South Asian background, but very little for males of South Asian background or people from other ethnic groups. Differences in household composition are therefore unlikely to be the main explanation of ethnic inequalities in COVID-19 outcomes, but may make an important contribution for some specific population subgroups, and may therefore be taken into account when prioritising vaccination. Relevant public health interventions (such as the provision of free accommodation to assist with self-isolation) should be considered to mitigate risks of infection spread within a household. Ensuring such interventions are accessible to communities where multi-generational households are highly prevalent (such as South Asian women) may be warranted.

## Introduction

People of ethnic minority background in the UK and the USA have been disproportionately affected by the Coronavirus disease 2019 (COVID-19) [1, 2, 3, 4, 5] compared to the White population, particularly Black and South Asian groups. Whilst several studies have investigated whether adjusting for socio-demographic and economic factors and medical history reduces the estimated difference in risk of mortality and hospitalisation [6, 7, 8], the reasons for the differences in the risk of experiencing harms from COVID-19 are still being explored.

One important driver of these ethnic inequalities may be differences in household structure between ethnic groups. Household composition varies substantially between ethnic groups, with some ethnic minority populations more likely to live in large, multi-generational households [9]. While living in multi-generational households is associated with increased social capital [10], which could have beneficial health effects [11], it may also increase the risk of potential viral transmission [12, 13]. For older people, who are at greater risk of experiencing severe complications if infected, residing with younger people may represent an increase in exposure to infection, which could lead to an increased risk of hospitalisation and mortality from COVID-19. To the best of our knowledge, no study has yet examined whether the difference in household composition partly explains the elevated risk of COVID-19 mortality in ethnic minority groups.

In this study, we examined the relationship between household composition and COVID-19 mortality risk amongst older adults (≥65 years) in England, with a focus on multi-generational households (older adults living with younger adults or dependent children). We then investigated how the propensity to live in a multi-generation household varies across ethnic groups, and whether this heterogeneity contributes to the raised risk of COVID-19 mortality amongst ethnic minority groups compared to the White population.

## Methods

### Data

This retrospective cohort study was based on the 2011 Census of England linked to mortality registration data and Hospital Episodes Statistics (2017 – 2019). The study population included all usual residents of England aged 65 years or over in 2020, who had been enumerated in private households at the time of the 2011 Census (27 March 2011), had not moved to a care home by 2019 (identified by linking to the NHS Patient Register) and were still alive on 2 March 2020. We further excluded individuals who entered the UK in the year before Census due to their higher propensity to leave the UK prior to the study period, and those aged over 100 years at the time of the Census. Our study population consisted of 10,078,568 individuals aged 65 years or over at 2 March 2020 (See Supplementary Table 1 details on the number of participants at each stage of the sample selection).

To adjust for out-migration, we applied weights reflecting the probability of having remained in the country until March 2020 after being enumerated in March 2011, based on data from the NHS Patient Register and the International Passenger Survey (IPS). Further information on the data has already been published [6]. All the variables used in the analysis, including their definitions and sources, are detailed in Table 1.

**Table 1.**
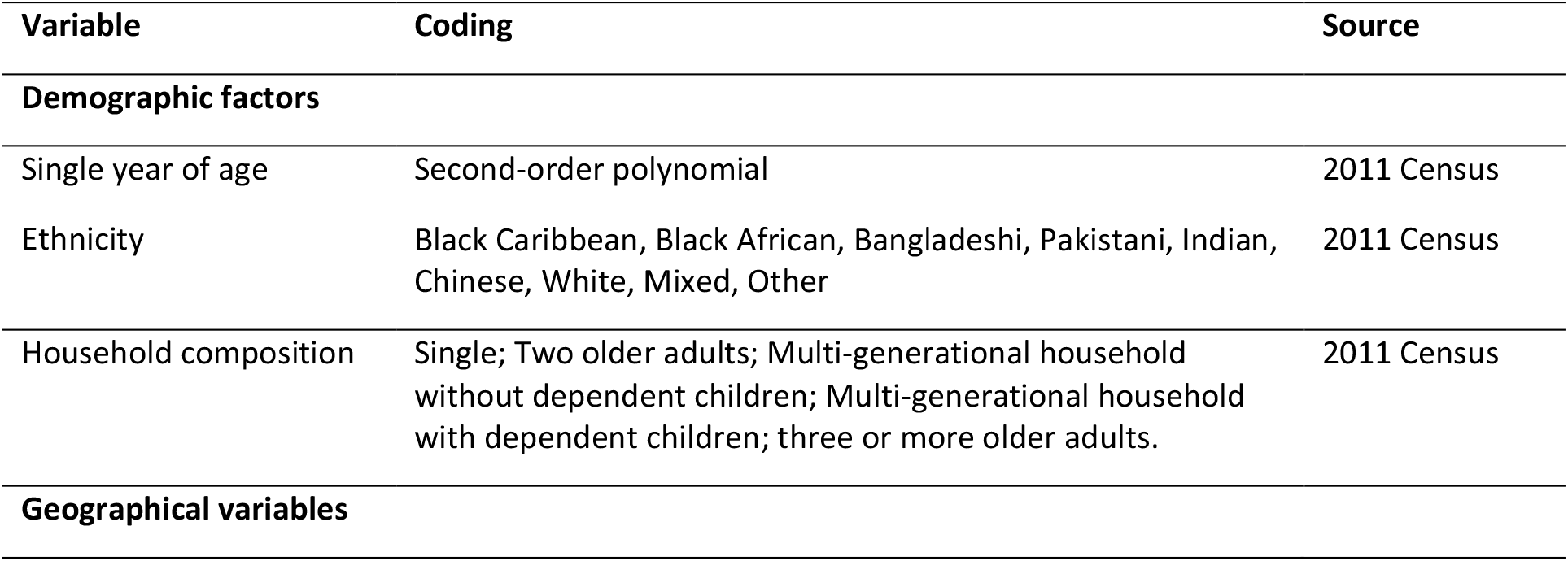
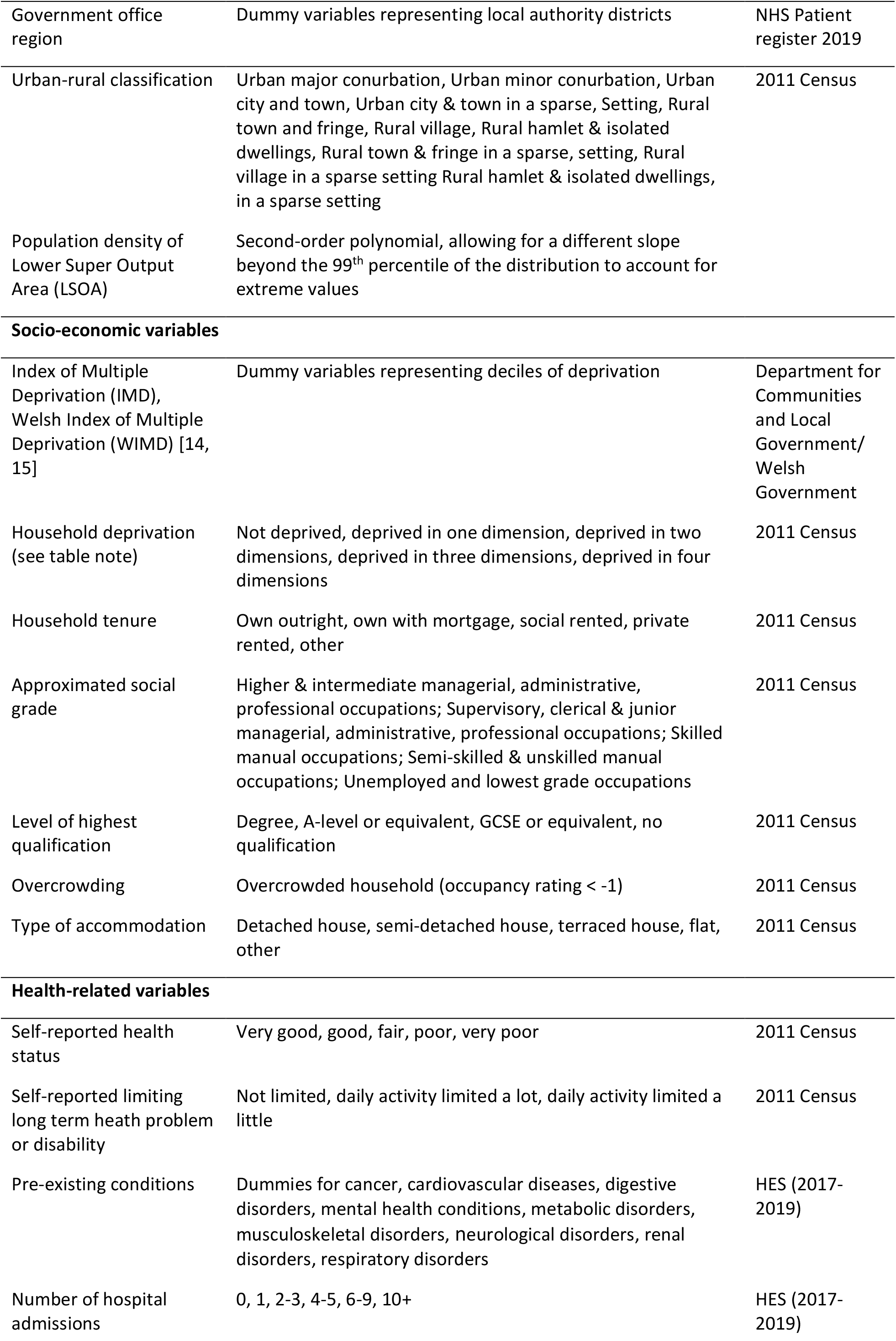
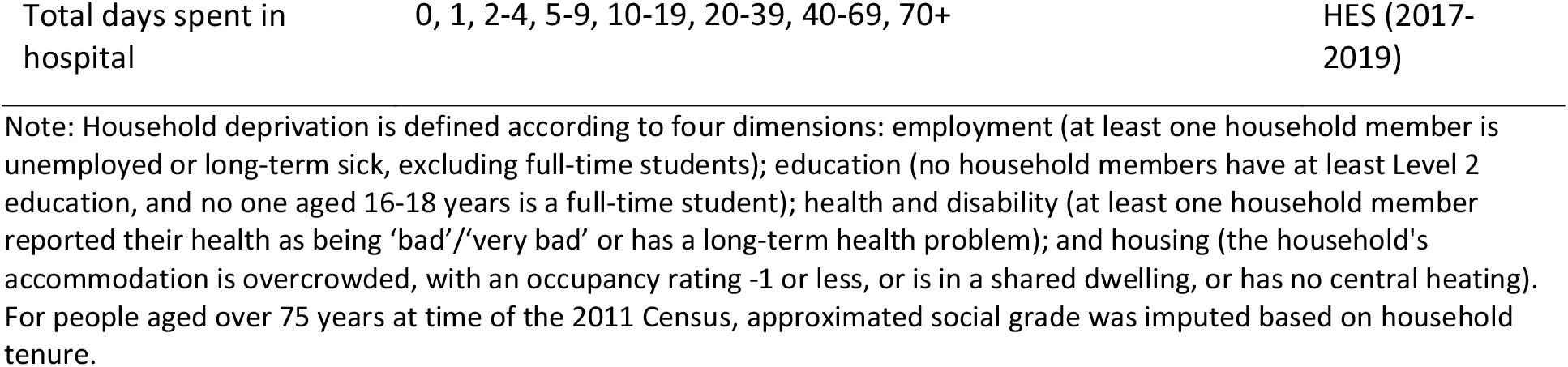
Variables used for the statistical modelling.

### Outcome and exposure

Deaths involving COVID-19 included those with an underlying cause, or any mention, of International Statistical Classification of Diseases and Related Health Problems 10^th^ Revision (ICD-10) codes U07.1 (COVID-19, virus identified) or U07.2 (COVID-19, virus not identified). We analysed deaths that occurred between 2 March 2020 and 28 July 2020, registered by 24 August 2020, which corresponds to the deaths that occurred during the first COVID-19 wave.

Household composition in 2020 was derived based the household composition at the time of the Census. We excluded people who died between 27 March 2011 and 1 March 2020 or had moved to a care home by 2019. To mitigate measurement error, we removed people aged 10 to 24 at the time of the Census because they are more likely to have moved out in 2020. We defined a multi-generational household to be a household in which someone aged 65 years or over on 2 March in 2020 co-resided with at least one other adult aged more than 20 years younger, or with at least one child. Our household composition variable classified households in five categories: Single; Two older adults; Multi-generational household without dependent children; Multi-generational household with dependent children; three or more older adults. As sensitivity analyses, we removed people aged 10 to 19 instead of 10 to 24).We also defined a multi-generational household to be one in which someone aged 65 years or over in 2020 co-resided with at least one other adult aged more than 15 years (instead of 20 years) younger. We also used longitudinal data from the English Longitudinal Study of Ageing (ELSA) to estimate change in household composition amongst adults aged 65 or over between 2008-09 and 2016-2017 (see the Technical Appendix for more details).

In the mediation analysis, the exposure was self-reported ethnic affiliation based on a nine-group classification (see Table 1). The two mediators were binary variables for living in a multi-generational household with or without children.

### Covariates

Demographic factors, geographical variables, socio-economic characteristics and measures of pre-pandemic health are listed in Table 1. These covariates were generally considered to be confounders of the relationship between household composition and COVID-19 mortality risk, and mediators of the ethnicity-mortality relationship (See Figure 1).

**Figure 1.**
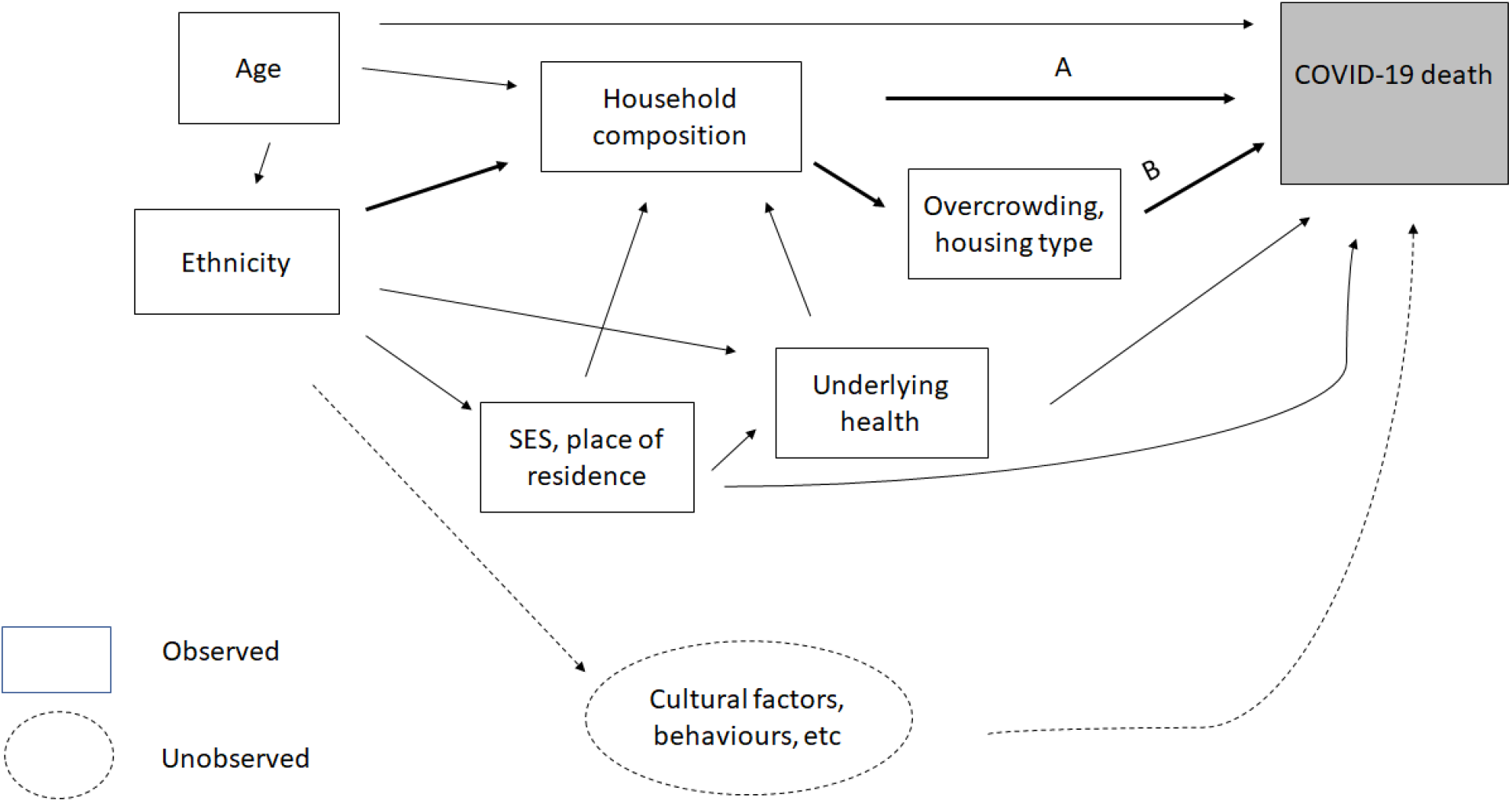
Directed Acyclic Graphs summarising the relationship between ethnicity, household composition and COVID-19 mortality. Note: When analysing whether household composition directly affects the risk of COVID-19 death, our effect of interest is *A*. In the mediation analysis, where we estimate the proportion of the ethnic disparity in COVID that is explained by living in a multi-generational household, the effects of interest are *A*+*B*

### Statistical analyses

We calculated age-standardised mortality rates (ASMRs) stratified by household composition and sex, separately for COVID-19 related deaths and other deaths. The ASMRs were standardised to the 2013 European Standard Population and can be interpreted as deaths per 100,000 of the population at risk during the analysis period.

We estimated Cox proportional hazard models to assess whether the risk of COVID-19-related death varies by household type (using living with one other older adult with as the reference category) after adjusting for the geographical factors, socio-economic characteristics and measures of health listed in Table 1. These factors could confound the relationship between household composition and COVID-19-related mortality, as shown in Figure 1. We estimated separate models for males and females, as the risk of death involving COVID19 differs markedly by sex [7]. When fitting the Cox models, we included all individuals who died during the analysis period and a weighted random sample of those who did not (5% of White people, and 20% amongst ethnic minority groups), and applied case weights to reflect the original population totals.

We conducted a causal mediation analysis [16] to estimate the proportion of excess risk in ethnic minority groups which is attributable to living in a multigenerational household. As a measure of inequality in COVID-19 mortality, ethnicity-specific odds ratios for COVID-19 mortality were estimated using logistic regression models, fitted to males and females separately and adjusting solely for age in the baseline model. The proportion of the difference in COVID-19 mortality rates between ethnic groups mediated by living in a multi-generational household was then estimated as the Average Causal Mediated Effect (ACME) as a proportion of the age-adjusted difference in the probability of COVID-19 mortality, using a non-parametric approach [17] (see the Technical Appendix for more details). The mediator models and the full outcome model were adjusted for geographical factors (region, population density, urban/rural classification), socio-economic characteristics (IMD decile, educational attainment, social grade, household tenancy) and health (self-reported health and disability from the Census, pre-existing conditions based on hospital contacts) but not for overcrowding or housing type, as these are likely to be consequences of living in a multi-generational household rather than confounding factors (See Figure 1). Confidence intervals were obtained via bootstrapping, using 500 replications. All statistical analyses were performed using R version 3.5.

## Results

### Characteristics of the study population

Characteristics of the study population are reported in Supplementary Table 2 in the Appendix. In our study population of 10,078,568 individuals in England aged 65 years or over who were not in a care home in 2019 and were still alive on 2 March 2020, just over half (53.9%) were female, the mean age was 75.2 years, and 93.9% reported being from a White ethnic background (Table 2). Over the outcome period (2 March 2020 to 28 July 2020), 27,989 (0.28%) died of COVID-19, and 123,551 (1.2%) died of other causes.

**Table 2.** Hazard ratios for COVID-19 related death for older adults (aged ≥65 years) in England, compared to living in a household with one other older adult, stratified by sex.

|  | Men |  | Women |  |
| --- | --- | --- | --- | --- |
|  | Age-adjusted | Fully-adjusted | Age-adjusted | Fully-adjusted |
| Single | 1.405 | 1.186 | 1.288 | 1.115 |
|  | [1.358 - 1.453] | [1.145 - 1.228] | [1.234 - 1.344] | [1.066 - 1.165] |
| Multi-generational household without children | 1.247<br>[1.181 - 1.318] | 1.027<br>[0.971 - 1.087] | 1.425<br>[1.319 - 1.539] | 1.215<br>[1.122 - 1.317] |
| Multi-generational household with children | 1.629<br>[1.464-1.812] | 1.128<br>[1.007 - 1.265] | 1.852<br>[1.622 - 2.114] | 1.169<br>[1.010 - 1.352] |
| 3+ older adults | 0.852<br>[0.695 - 1.046] | 0.900<br>[0.733 - 1.106] | 0.724<br>[0.556 - 0.944] | 0.824<br>[0.632 - 1.075] |
| Observations (unweighted) | 432,894 |  | 489,192 |  |
| Concordance | 0.7432 | 0.8284 | 0.7588 | 0.8551 |

Note: Hazard ratios compared to living in a household with one other older adult. Fully adjusted Cox regression models include geographical factors (region, population density, urban/rural classification), ethnicity, socio-economic characteristics (IMD decile, household deprivation, educational attainment, social grade, household tenancy), health (self-reported health and disability from the Census, pre-existing conditions based on hospital contacts, number of hospital admissions, total days spent in hospital), a measure for overcrowding, and property type.

Compared with older adults living with one other older adult (*n* = 5,538,963), people living by themselves (*n* = 3,287,395) had a higher mean age, were more likely to be female, and tended to be more deprived. Older people living in a multi-generational household without dependent children (*n* = 987,306) and with dependent children (n = 199,112) were on average younger and were more likely to be from an ethnic minority group, live in London and large urban conurbations, and tended to be more deprived than older people living with another older adult.

*Figure 2* shows that household composition varied substantially between ethnic groups. Among older people, just over 10% of those of White background lived in a multi-generational household, compared to over half of Bangladeshi or Pakistani background (58.7% and 58.8% respectively) and 45.8% of Indian background. The patterns were similar for males and females, although a larger proportion of females live by themselves (See Supplementary Figure 1).

**Figure 2.**
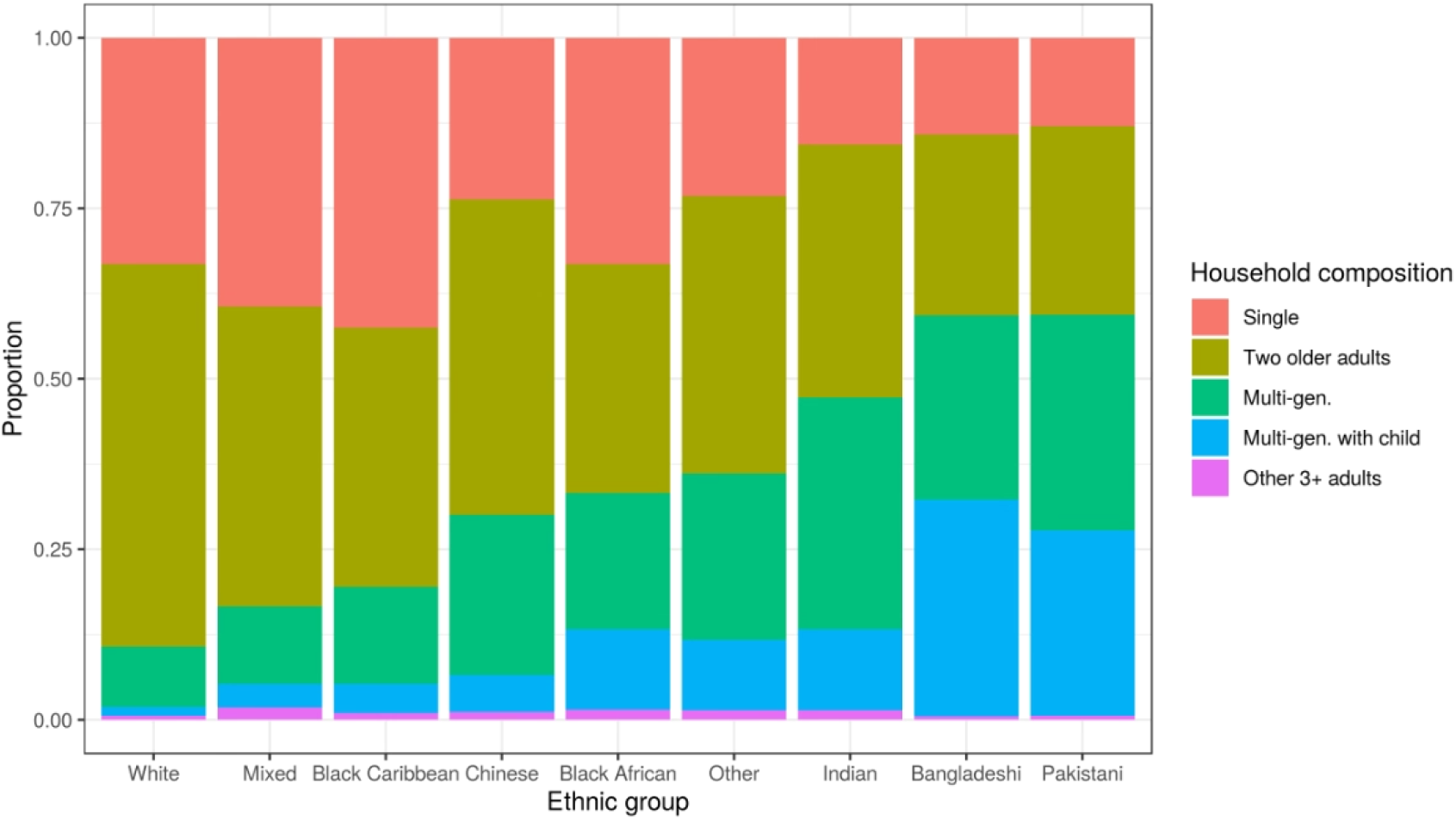
Household composition by ethnic group for people in England aged ≥65 years. Note: Linked 2011 Census and mortality registration data for people in England aged ≥65 years, excluding those living in a care home in 2019. The number of adults in the household was calculated as the number of people aged ≥25 years who lived in the household at the time of the Census, minus those who died between 27 March 2011 and 1 March 2020.

### Household composition and death involving COVID-19

Older people living by themselves were more likely to have died from COVID-19 over the study period than those living with another adult. For males, the ASMRs were 457 [95% confidence interval (CI): 446-469] and 330 [322-337] per 100,000 of the population for those living by themselves and those living with another adult, respectively (Figure 2, Panel A). For females, the ASMRs were 217 [212-223] and 167 [161-173] per 100,000, respectively. A similar pattern is observed for deaths from other causes (Figure 2, Panel B).

There was a positive association between the risk of COVID-19 death and living a in multi-generational household. Both older males and females living a multi-generational household without school-age children were more likely to die from COVID -19 than older people living with another older adult (ASMR 394 [373-416] per 100,000 for males, 228 [211-245] for females), with the risk of death being greater still if there were children in the household (ASMR 499 [443-555] per 100,000 for males, 287 [249-325] per 100,000 for females). The risk of COVID-19 mortality was higher in males than that in females across all the household compositions. There was no clear relationship between living in a multi-generational household and mortality from other causes.

Adjusting for individual- and household-level characteristics (including age, geographical factors, socio-economic characteristics and measures of pre-pandemic health) reduced the estimated differences in COVID-19 mortality rates between older adults living in different types households (Table 2). However, even after adjusting for these characteristics, living in a multi-generational household, especially with children, remained associated with an increased risk of COVID-19-related death. Compared to living with another older adult aged 65 years or above, the rate of COVID-19-related death was 1.22 [1.12 - 1.32] and 1.17 [1.01 - 1.35] times greater for older females living in a multi-generational household without and with children, respectively. For older males, after adjusting for individual and household characteristics, there was no evidence of an association between living in a multi-generational household without children and the risk of COVID-19 mortality (HR: 1.03 [0.97-1.09]). However, living in a multi-generational household with children was associated with a 1.13 [1.01-1.27] times greater risk of COVID-19 related death. The rate of COVID-19 related death was also 1.19 [1.15-1.23] times greater for older males living alone than for those living with another older adult, and 1.13 [1.08 -1.18] times greater for older females. The results were similar in the sensitivity analyses using different definitions of household composition (See Supplementary Table 3).

### Living in a multi-generational household as a mediator for the disparity in COVID-19 death between ethnic groups

Figure 4 shows the decomposition of the age-adjusted odds ratios of COVID-19 death for ethnic minority groups compared to those of white ethnic group into three parts: (i) the part explained by living in a multi-generational household (in red); (ii) the part explained by other individual and household characteristics, such as geographical factors, socio-economic factors and pre-pandemic health (in green); and (iii) a residual component that is not explained by our model (in blue).

**Figure 3.**
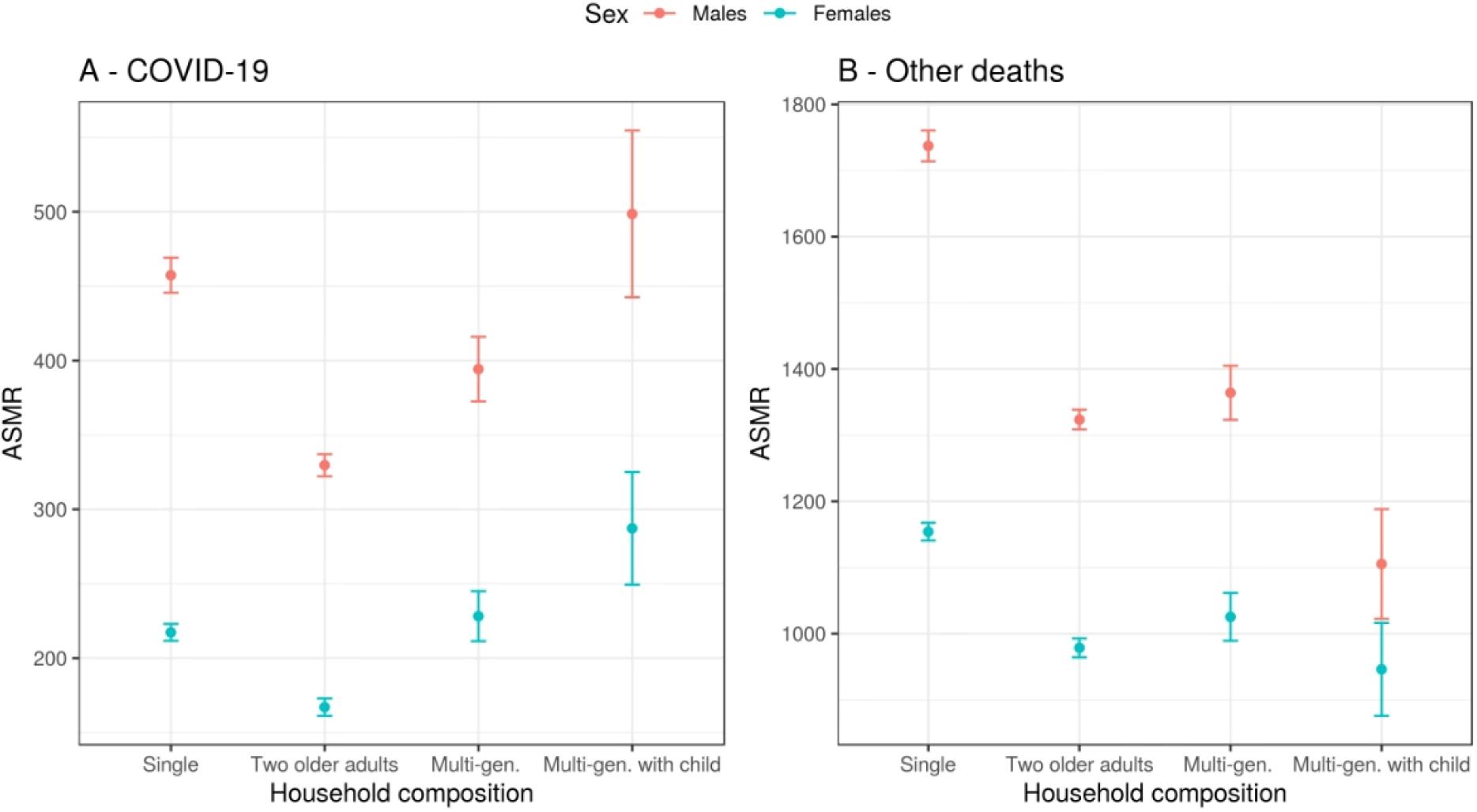
Age-standardised mortality rates per 100,000 adults aged 65 years or over, stratified by sex and household composition. Note: Deaths occurring between 2 March 2020 and 28 July 2020. 95% Confidence intervals are reported. Mortality rates are standardised to the 2013 European Standard Population.

**Figure 4.**
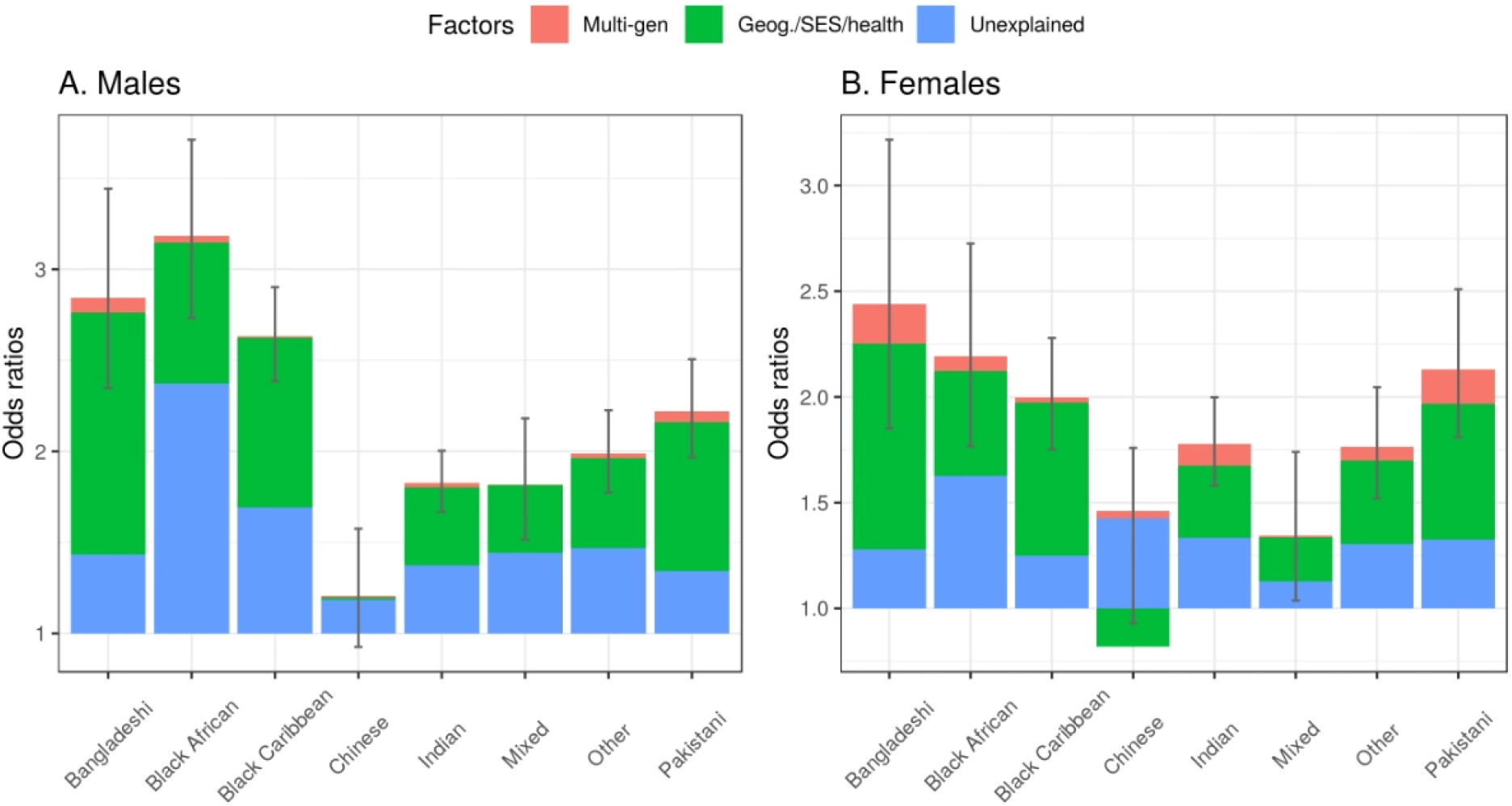
Decomposition of odds ratios for COVID-19 mortality amongst older adults (aged ≥65 years) across ethnic groups, stratified by sex. Note: The overall height of the bar corresponds to the odds ratio (OR), relative to the White population, based on a logistic regression model adjusted for age. Error bars are 95% confidence intervals. The proportion of the age-adjusted ORs explained by living in a multi-generational household were calculated through a mediation analysis. The unexplained part corresponds to the ORs from a model adjusted for age, geographical factors (region, population density, urban/rural classification), socio-economic characteristics (IMD decile, household deprivation, educational attainment, social grade, household tenancy), health: (self-reported health and disability from the Census, pre-existing conditions based on hospital contacts, number of hospital admissions, total days spent in hospital)

Among people aged 65 years or over, those from all ethnic minority groups except Chinese were at greater risk of COVID-19 related death than those from the White population. The odds of COVID-19 death were 3.1 [2.7-3.6] times greater for older Black African males and 2.2 [1.8-2.7] times greater for Black African females than for males and females from White ethnic group. The odds of death were also notably greater for people of Bangladeshi, Black Caribbean or Pakistani ethnic background than the White population, with odds ratios of 2.8 [2.3-3.4], 2.6 [2.4-2.9], and 2.2 [1.9-2.5], respectively, for males and 2.5 [1.9-3.3], 2.0 [1.7-2.3] and 2.2 [1.8-2.7], respectively, for females.

Living in a multi-generational household did not explain much of the difference in COVID-19 mortality rates amongst older males. However, it did explain a substantial part of the difference between older females of South Asian background and White older females. For older females of Pakistani background, living in a multi-generational household accounted for 13.8% [3.4%-23.9%] of the difference in COVID-19 mortality rates compared to White females; for females of Bangladeshi or Indian background, it accounted for 12.3% [7.3%-19.5%] and 12.2% [5.7% - 23.0%] of the difference in mortality rates, respectively (See Supplementary Table 4 in appendix for full results). The results were similar in the sensitivity analyses using different definitions of household composition (See Supplementary Table 5).

## Discussion

### Principal findings

This paper makes two contributions to the research on COVID-19. First, we find that, among older adults, household composition is associated with COVID-19 mortality, even after adjusting for a range of socio-demographic factors and measures of health. Our results indicate that compared to those living in a two older adult household, older adults, especially females, living in a multi-generational household are at greater risk of COVID-19 death. Living alone is also associated with elevated COVID-19 mortality. Second, we find that living in a multi-generational household explains between 10% and 15% of the excess COVID-19 mortality risk for females of South Asian background, but very little for males or people from other ethnic groups.

### Comparison with related studies

Our results are consistent with emerging evidence that household size is associated with the risk of infection [18, 19], and that older adults tend to be at greater risk of household transmission [20, 21]. Older people living in large household tend to live in multi-generational households, co-habiting with younger adults and children. There is some evidence that, amongst older adults, living with dependent children is not strongly associated with the risk of COVID-19 infection or adverse outcomes [22]. Whilst our results indicate that older adults living in a multi-generational household are at greater risk of COVID-19 death compared to those living with another older adult, we find little difference in risk between older people living in a multi-generational household with or without young children.

Several studies have analysed ethnic differences in COVID-19 infection and mortality [3, 7, 6, 8, 4]. Although we focus on older adults only, we find that almost all ethnic minority groups were at higher risk of COVID-19 deaths compared to the White population, and that the differences were attenuated once we adjusted for a range of geographical factors, socio-demographic characteristics and comorbidities. We improve the existing evidence on ethnic inequalities in COVID-19 mortality by using a causal mediation approach to quantify the importance of living in a multi-generational household.

### Mechanisms

Our results suggest that older people are placed at increased exposure to infection by living with younger adults rather than young children. After adjusting for confounding factors, we find that the risk of COVID-19 death is similar amongst older adults living in a household with young children and those living in a household with younger adults only. The increased risk is likely to be driven by co-residing with younger adults, who have a higher risk of infection than older people [19]. Younger adults are likely to be at increased risk of exposure because of work, as evidence suggests that in England people who are working were at greater risk of infection compared to people not in employment, especially if they were working in patient or client-facing occupations [19, 23, 24].

Older adults living by themselves were also found to be at greater risk of COVID-19 death than those living with another older adult. During the COVID-19 pandemic, older people living alone were more likely to have received help from carers, including informal helpers, than people living with another older adult [25]. These frequent contacts with people from different households could increase the risk of being exposed to the virus.

We find that living in a multi-generational household explains between 10% and 15% of the excess COVID-19 mortality risk for females of South Asian background, but very little for males, despite a similar proportion of them living in a multi-generational household. Women spend more time at home than men and still do the majority of unpaid housework [26], which could increase the risk of household transmission.

### Strengths and limitations

The primary strength of our study lies in the use of a unique linked population-level dataset which combine the 2011 Census with death registration data and hospital records. Unlike data based solely on health records, our study dataset contains a broad range of information on demographic, socio-economic, and household characteristics, including occupation. Unlike sample survey data, it contains millions of observations covering the entire population of interest, allowing us to examine both the association between household composition and COVID-19 mortality and also whether living in a multi-generational household explains some of the disparity in COVID-19 mortality between ethnic groups. We were able to examine differences between disaggregated ethnic minority groupings rather than high-level categories of South Asian, Black, and Other.

The main limitation of our study is that household composition is likely to be imprecisely measured. Whilst household composition is based on a detailed and accurate measurement taken in 2011, we could only identify changes since then due to death of household members or a move to a care home. Whist we took several steps to limit the measurement error, such as focusing on older adults, including only adults aged 25 or over and children aged 0 to 9 at the time of the census in our definition of household composition, our household composition measure may not reflect current living circumstances of everybody in our population of interest. To mitigate concerns about measurement error, we showed that our results are robust to using different definitions of household composition. Nonetheless, measurement error is likely to attenuate the explanatory power of household composition in our models. In addition, while we have used a causal mediation approach, our analysis remains based on observational data and therefore residual confounding is likely. Another limitation is that our statistical approach assumes that the effect of living in different types of household composition is the same across ethnic groups.

### Conclusions

Older adults living in multi-generational households are at elevated risk of experiencing harms from COVID-19 compared to older adults living with people of the same age. However, there has been little focus on implementing effective interventions (such as creating plans to effectively isolate and improving ventilation within the home) to reduce transmission risk within the household [26].

Relevant public health interventions should be directed at communities where multi-generational households are highly prevalent. Living in a multi-generational household explains some of the excess COVID-19 mortality risk for females of South Asian background, but very little for males or people from other ethnic groups. Further research is needed to explain the difference in COVID-19 mortality between ethnic groups.

## Data Availability

Under the provisions of the Statistics and Registration Service Act 2007, the linked 2011 Census data used in this study are not permitted to be shared.

## Footnotes

### Funding

This research was funded by the Office for National Statistics. VN is also funded by Health Data Research UK (HDR-UK). HDR-UK is an initiative funded by the UK Research and Innovation, Department of Health and Social Care (England) and the devolved administrations, and leading medical research charities. SVK acknowledges funding from a NRS Senior Clinical Fellowship (SCAF/15/02), the Medical Research Council (MC_UU_12017/13) and the Scottish Government Chief Scientist Office (SPHSU13).”

### Ethics approval

Following assessment using the National Statistician’s Data Ethics Advisory Committee (NSDEC)’s tool, we engaged with the UK Statistics Authority Data Ethics team and it was decided that ethical approval was not required. This is standard practice for analysing national Census data.

### Author contributors

VN, NI, DA, CL and KK contributed to the study conceptualisation and design. VN lead the preparation of the study data and performed the statistical analyses. AS assisted with the preparation of the data. All authors contributed to interpretation of the results. VN drafted the manuscript. DA, NI, CG, VK, RM, MA, PG, AJ, AB, SG and KK contributed to the critical revision of the manuscript. All authors approved the final manuscript. VN is the guarantor for the study. The corresponding author attests that all listed authors meet authorship criteria and that no others meeting the criteria have been omitted.

### Conflict of interest statement

Prof. Khunti is a member of the Independent Scientific Advisory Group for Emergencies (SAGE), a Trustee of the SAHF, Director of the NIHR Applied Research Collaboration (ARC) East Midlands, and Director of the Centre for Black and Minority Ethnic (BME) Health. The other authors declare no competing interests.

### Appendix

#### Change in household composition in the English Longitudinal Study of Ageing

Using a measure of household composition based on outdated information from the 2011 Census may introduce some measurement error. To quantify this measurement error, we used data from wave 4 (2008-2009) and wave 8 (2016-2017) of the English Longitudinal Study of Ageing (ELSA). We identified whether respondents. We used core members of the study who were interviewed in wave 4 and wave 8 of ELSA, were aged 65 years or older in the year of their wave 4 interview and lived in a private household. We identified those who lived with their adult children (aged 25 or over in wave 4) in wave 4 and in wave 8 and estimated the proportion who had a different co-residence pattern in wave 4 and 8. the mean period between a Wave 4 and Wave 8 interview was 7.98 years (sd = 0.07 years).

Most people (93.1%) had the same co-residence pattern in 2008-2009 and 2016-2017. Nearly all (96.6%) of people who did not live with an adult child in wave 4 did not live with an adult child in wave 8. The majority (61.2%) of people who lived with an adult child in wave 4 also lived with an adult child in wave 8.

#### Estimating the Average Causal Mediated Effect

The Average Causal Mediated Effect (ACME) of living in a multi-generational household for ethnic group k compared to the white group, δ(*k*) is estimated as:

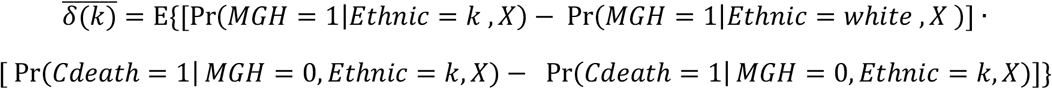

Where *MGH* indicates if individual i lives in a multi-generational household, *Ethnic* is the ethnic group and X is a vector of factors likely to confound the relationship between the mediator and the outcome. X includes geographical factors (region, population density, urban/rural classification), socio-economic characteristics (IMD decile, household deprivation, educational attainment, social grade, household tenancy), health: (self-reported health and disability from the Census, pre-existing conditions based on hospital contacts, number of hospital admissions, total days spent in hospital).

To estimate each component 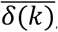, we use predicted probabilities based on logistic regression models.

The total estimated difference in probability of COVID-19 death between ethnic group k and the White ethnic group is given by:

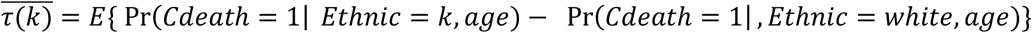

The proportion of the difference in the probability to die from COVID-19 between ethnic groups that is mediated by living in a multi-generational household is given by the ACME, 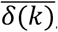, as a proportion of the total effect 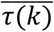. We then used this proportion to decompose the age-adjusted odds ratios.

**Supplementary Table 1:**
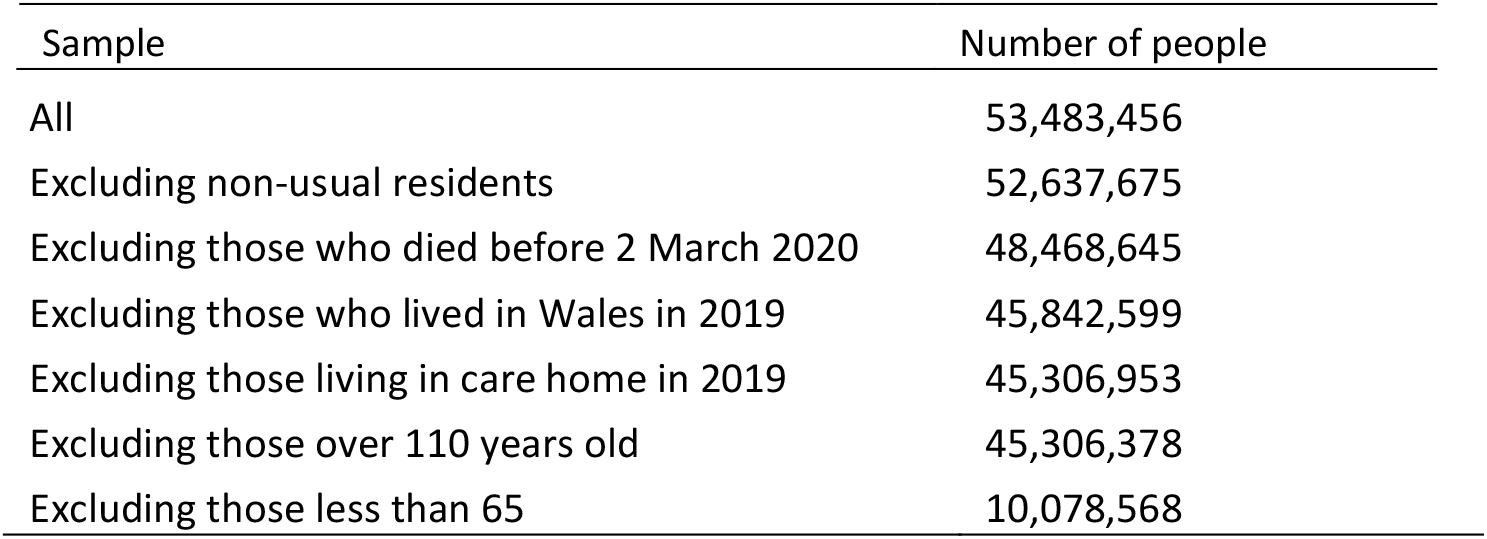
Sample selection and number of participants.

**Supplementary Table 2:**
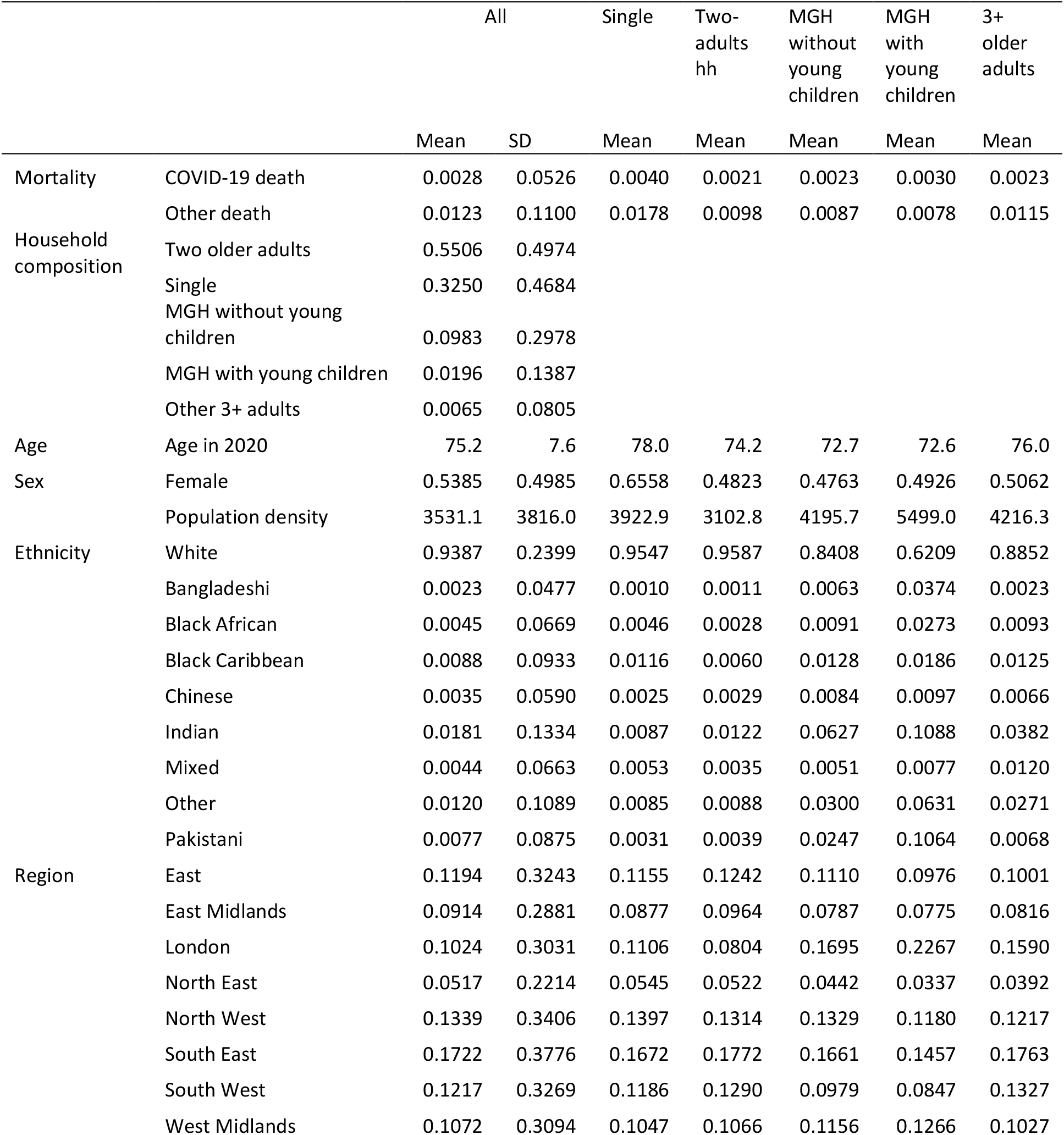
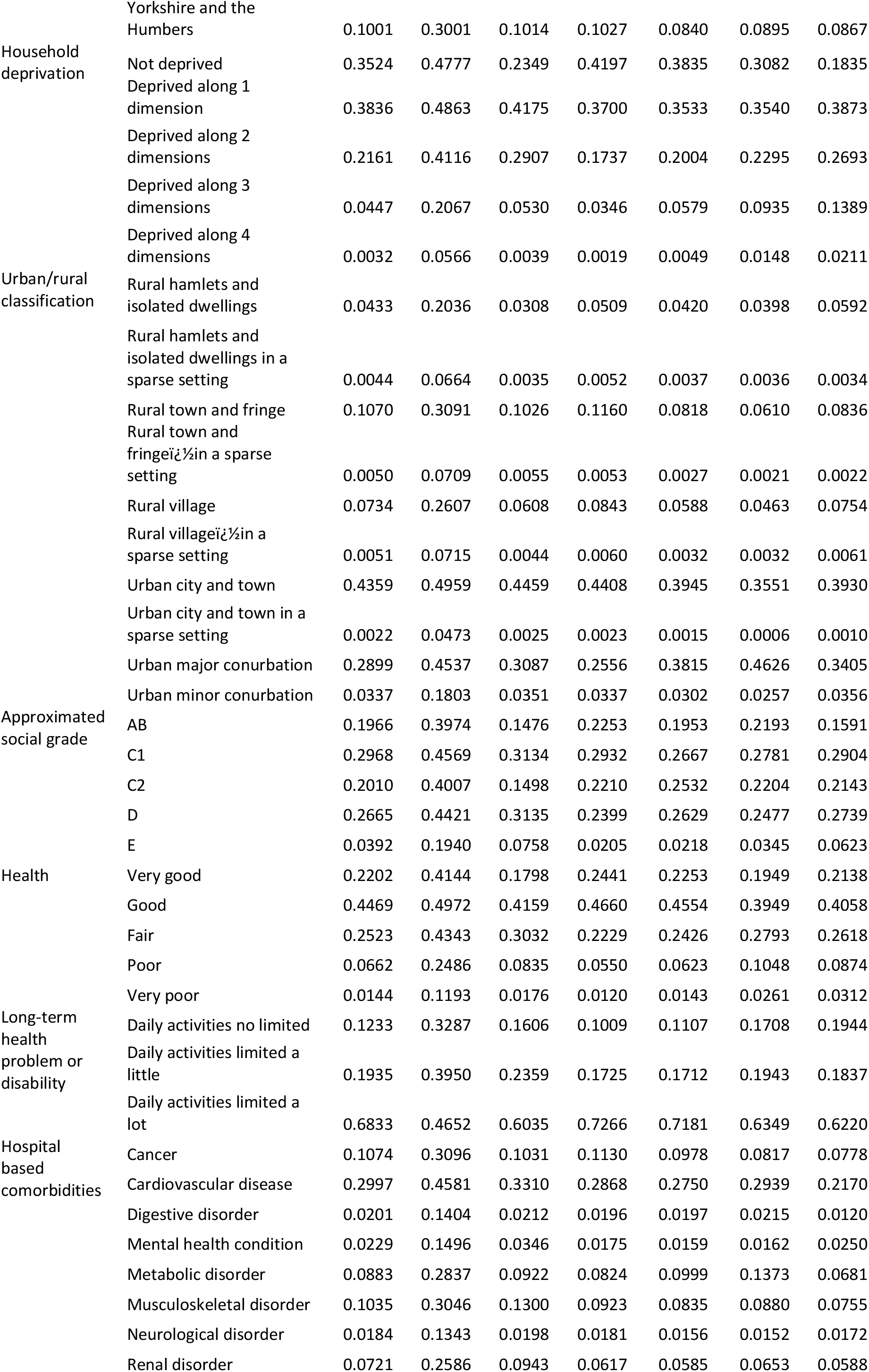
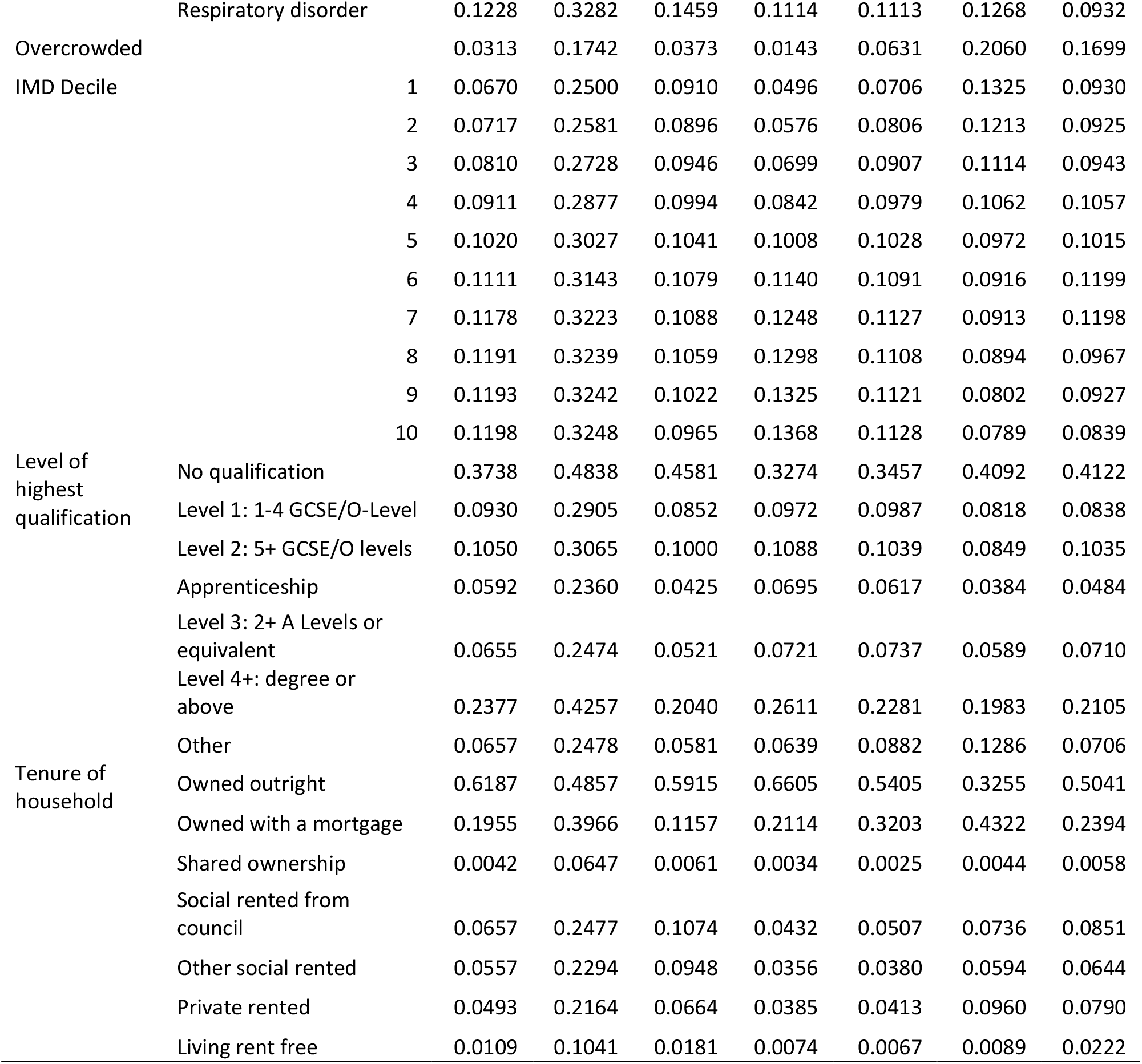
Distributions of study variables, stratified by household composition:

**Supplementary Figure 1.**
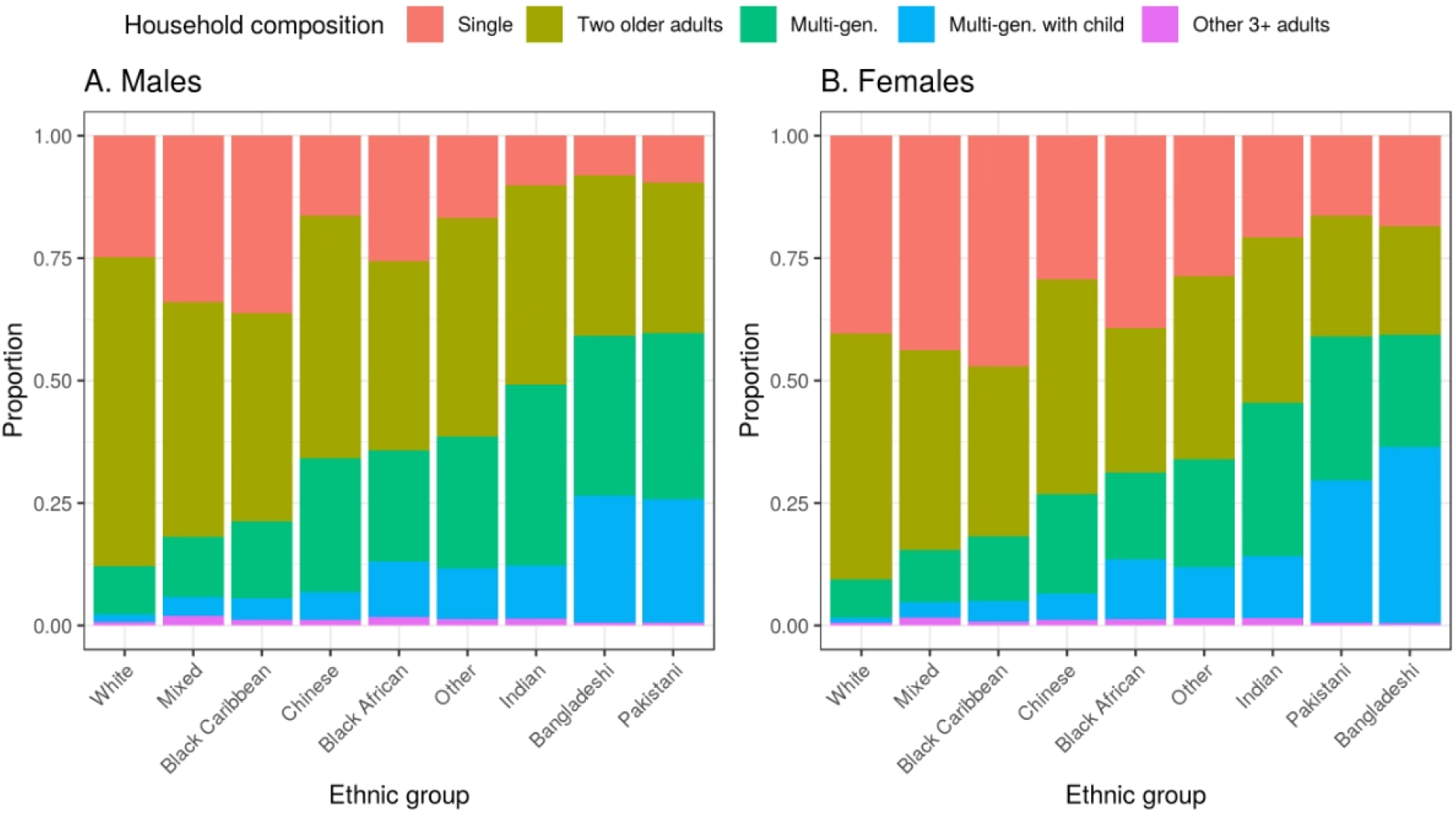
Household composition by ethnic group for people in England aged ≥65 years, stratified by sex. Linked 2011 Census and mortality registration data for people in England aged ≥65 years, excluding those living in a care home in 2019. The number of adults in the household was calculated as the number of people aged ≥16 years who lived in the household at the time of the Census, minus those who died between 27 March 2011 and 1 March 2020.

**Supplementary Table 3:** Hazard ratios for COVID-19 related death for older adults (aged ≥65 years) in England, compared to living in a household with one other older adult, using different definitions of household composition, stratified by sex.

|  | Men |  | Women |  |
| --- | --- | --- | --- | --- |
|  | Def. A1 | Def. A2 | Def. A1 | Def. A2 |
| Single | 1.164<br>[1.123 - 1.206] | 1.164<br>[1.124 - 1.207] | 1.12<br>[1.071 - 1.170] | 1.12<br>[1.071 - 1.171] |
| Multi-generational household without children | 1.032<br>[0.975 - 1.092] | 1.027<br>[0.974 - 1.083] | 1.207<br>[1.115 - 1.308] | 1.226<br>[1.135 - 1.323] |
| Multi-generational household with children | 1.129<br>[1.007-1.266] | 1.128<br>[1.006-1.266] | 1.165<br>[1.007 - 1.348] | 1.176<br>[1.016 - 1.361] |
| 3+ older adults | 0.873<br>[0.702 - 1.085] | 0.92<br>[0.749 - 1.131] | 0.761<br>[0.577 - 1.002] | 0.715<br>[0.537 -0.953] |
| Observations | 432,894 |  | 489,192 |  |

Note: Def A1: we derived household composition in 2020 based on the number of adults aged 20 years (instead of 25 years); Def A2: multi-generational household as household with someone aged 65 years or over in 2020 co-resided with at least one other adult aged more than 15 years (instead of 20 years) younger. Hazard ratios compared to living in a household with one other older adult. Fully adjusted Cox regression models include geographical factors (region, population density, urban/rural classification), ethnicity, socio-economic characteristics (IMD decile, household deprivation, educational attainment, social grade, household tenancy), health (self-reported health and disability from the Census, pre-existing conditions based on hospital contacts, number of hospital admissions, total days spent in hospital), a measure for overcrowding, and property type.

**Supplementary Table 4:** Proportion of difference in COVID-19 mortality rates between ethnic groups mediated by living in a multi-generational household, stratified by sex.

|  | Multi-gen with young children |  | Multi-gen without young children |  | Multi-gen (total) |  |
| --- | --- | --- | --- | --- | --- | --- |
|  | Prop. | 95% CI | Prop. | 95% CI | Prop. | 95% CI |
| **Females** |  |  |  |  |  |  |
| Bangladeshi | 0.0856 | [0.0057 - 0.1738] | 0.0379 | [0.0195 - 0.0647] | 0.1234 | [0.0444 - 0.2155] |
| Black African | 0.0296 | [0.0020 - 0.0615] | 0.0261 | [0.0138 - 0.0414] | 0.0557 | [0.0254 - 0.0937] |
| Black Caribbean | 0.0083 | [0.0005 - 0.0169] | 0.0150 | [0.0078 - 0.0238] | 0.0233 | [0.0123 - 0.0368] |
| Chinese | 0.02750 | [-0.1331 - 0.2331] | 0.1011 | [-0.4773 - 0.6481] | 0.1286 | [-0.6403 - 0.8909] |
| Indian | 0.0371 | [0.0023 - 0.0743] | 0.0852 | [0.0465 - 0.1282] | 0.12223 | [0.0726 - 0.1834] |
| Mixed | 0.0107 | [-0.0128 - 0.0636] | 0.0100 | [-0.0137 - 0.0499] | 0.0207 | [-0.0339 - 0.1001] |
| Other | 0.0305 | [0.0018 - 0.0630] | 0.0489 | [0.0255 - 0.0764] | 0.0794 | [0.0430 - 0.1260] |
| Pakistani | 0.0745 | [0.0045 - 0.1519] | 0.0630 | [0.0343 - 0.0969] | 0.1375 | [0.0651 - 0.2230] |
| **Males** |  |  |  |  |  |  |
| Bangladeshi | 0.0361 | [-0.0062 - 0.0810] | 0.0085 | [-0.0150 - 0.0270] | 0.0446 | [-0.0070 - 0.0953] |
| Black African | 0.0125 | [-0.0024 - 0.0312] | 0.0038 | [-0.0074 - 0.0132] | 0.0163 | [-0.0035 - 0.0381] |
| Black Caribbean | 0.0033 | [-0.0007 - 0.0088] | 0.0019 | [-0.0035 - 0.0061] | 0.0051 | [-0.0015 - 0.0128] |
| Chinese | 0.0209 | [-0.2011 - 0.4050] | 0.0254 | [-0.1984 - 0.4917] | 0.0462 | [-0.3185 - 0.8844] |
| Indian | 0.0192 | [-0.0034 - 0.0449] | 0.0138 | [-0.0246 - 0.0421] | 0.0330 | [-0.0112 - 0.0704] |
| Mixed | 0.0033 | [-0.0006 - 0.0090] | 0.0007 | [-0.0016 - 0.0033] | 0.0040 | [-0.0008 - 0.0106] |
| Other | 0.0157 | [-0.0030 - 0.0369] | 0.0157 | [-0.0132 - 0.0239] | 0.0232 | [-0.0057 - 0.0521] |
| Pakistani | 0.0407 | [-0.0077 - 0.0926] | 0.0407 | [-0.0167 - 0.0300] | 0.0505 | [-0.0083 - 0.1085] |

Note: Proportion of difference in COVID-19 mortality between ethnic group mediated by living in a multi-generational household is estimated as the Average Causal Mediated Effect (ACME) as a proportion of the age-adjusted difference in the probability of COVID-19 death. The ACME is derived based on models that adjust for geographical factors (region, population density, urban/rural classification), socio-economic characteristics (IMD decile, household deprivation, educational attainment, social grade, household tenancy), and health: (self-reported health and disability from the Census, pre-existing conditions based on hospital contacts). 95% Confidence intervals are obtained via bootstrapping, using 500 replications.

**Supplementary Table 5:**
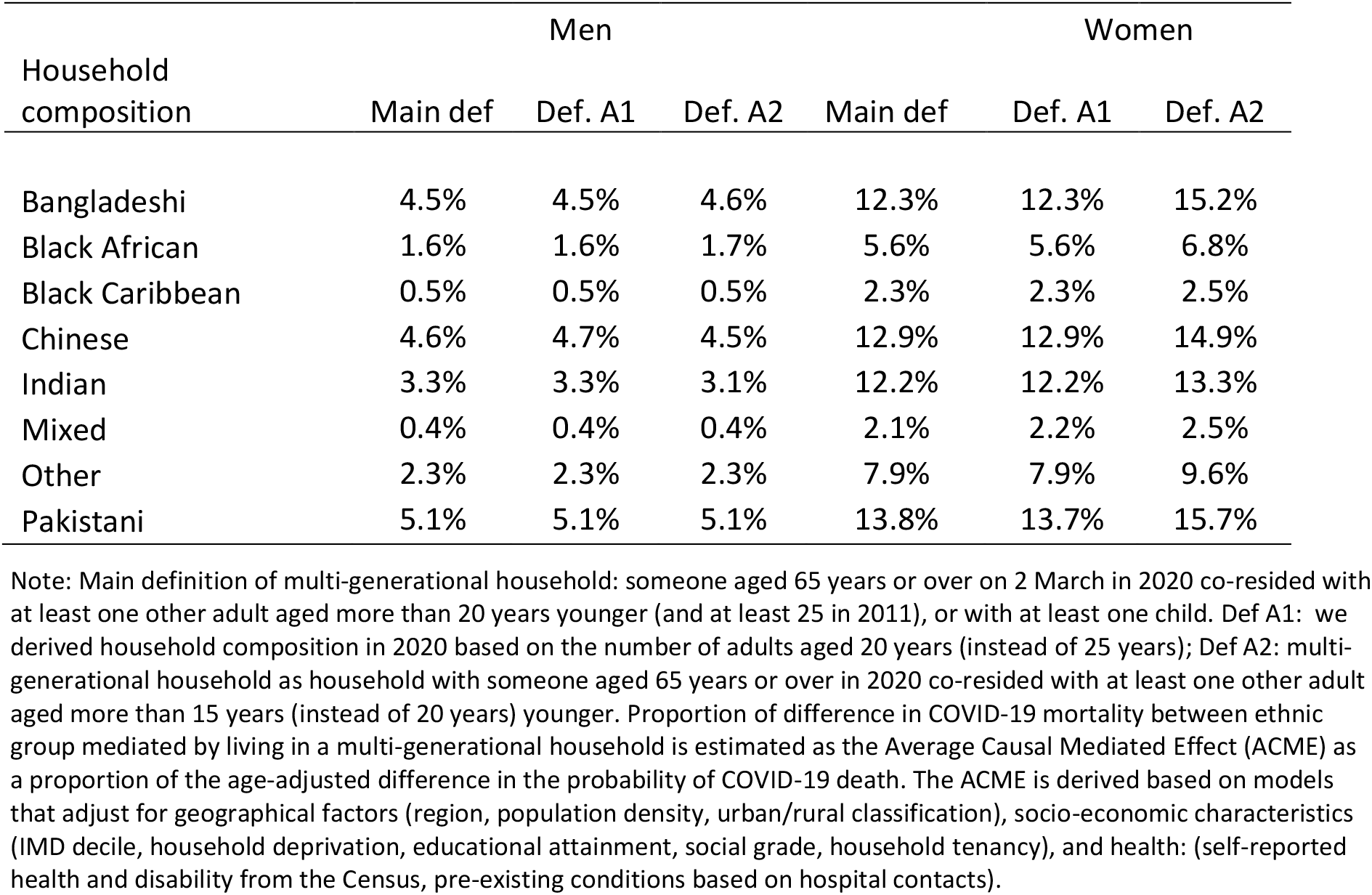
Proportion of difference in COVID-19 mortality rates between ethnic groups mediated by living in a multi-generational household using different definitions of household composition, stratified by sex.

| Household composition | Men |  |  | Women |  |  |
| --- | --- | --- | --- | --- | --- | --- |
|  | Main def | Def. A1 | Def. A2 | Main def | Def. A1 | Def. A2 |
| Bangladeshi | 4.5% | 4.5% | 4.6% | 12.3% | 12.3% | 15.2% |
| Black African | 1.6% | 1.6% | 1.7% | 5.6% | 5.6% | 6.8% |
| Black Caribbean | 0.5% | 0.5% | 0.5% | 2.3% | 2.3% | 2.5% |
| Chinese | 4.6% | 4.7% | 4.5% | 12.9% | 12.9% | 14.9% |
| Indian | 3.3% | 3.3% | 3.1% | 12.2% | 12.2% | 13.3% |
| Mixed | 0.4% | 0.4% | 0.4% | 2.1% | 2.2% | 2.5% |
| Other | 2.3% | 2.3% | 2.3% | 7.9% | 7.9% | 9.6% |
| Pakistani | 5.1% | 5.1% | 5.1% | 13.8% | 13.7% | 15.7% |

Note: Main definition of multi-generational household: someone aged 65 years or over on 2 March in 2020 co-resided with at least one other adult aged more than 20 years younger (and at least 25 in 2011), or with at least one child. Def A1: we derived household composition in 2020 based on the number of adults aged 20 years (instead of 25 years); Def A2: multi-generational household as household with someone aged 65 years or over in 2020 co-resided with at least one other adult aged more than 15 years (instead of 20 years) younger. Proportion of difference in COVID-19 mortality between ethnic group mediated by living in a multi-generational household is estimated as the Average Causal Mediated Effect (ACME) as a proportion of the age-adjusted difference in the probability of COVID-19 death. The ACME is derived based on models that adjust for geographical factors (region, population density, urban/rural classification), socio-economic characteristics (IMD decile, household deprivation, educational attainment, social grade, household tenancy), and health: (self-reported health and disability from the Census, pre-existing conditions based on hospital contacts).

